# Seizure cycles in paediatric epilepsy

**DOI:** 10.1101/2023.11.01.23297587

**Authors:** Hannah Kamitakahara, Philippa J. Karoly, Rachel E. Stirling, Dominique Eden, Ewan S. Nurse, Gabriel Dabscheck, Dean Freestone, Mark J. Cook, Wendyl D’Souza, Jodie Naim-Feil

## Abstract

Multiday cyclic patterns underlying the timing of seizures are well-established in adults with epilepsy and are critical to the development of seizure risk forecasting models. As cycles underpinning these models are yet to be explored in paediatric cohorts, the current study applies methods drawn from seizure risk forecasting to identify and compare multiday seizure cycles between paediatric and adult cohorts. This is followed by the first validation of personalised forecasts of seizure likelihood in a paediatric cohort. Multiday seizure cycles were extracted retrospectively from 325 (71 paediatric) electronic seizure diary users (more than 28 days of app use) with confirmed epilepsy. Cycles were grouped (k-means clustering), and seizure cycles quantified (synchronisation index), with significant cycles identified by Rayleigh test of periodicity (*p*<0.05). Wilcoxon rank-sum test assessed differences in prevalence and strength of cycle groups between paediatric and adult cohorts. 34,402 seizures (paediatric: *M*=101, *SD*=103, adult: *M*=107, *SD*=156) were analysed and seizure cycles were grouped according to circadian (0.5-1.5 days), about-weekly (2-12 days), about-fortnightly (13-22 days) and about-monthly (23-32 days) periodicities. Significant cycles were identified in each cycle group, with no differences in prevalence or cycle strength between paediatric and adult cohorts for any multiday cycle group. Similar effects were observed across clinical and demographic features (sex, epilepsy-type, medication). These multiday patterns formed the basis for cycle-based estimates of seizure likelihood. Receiver operating characteristic (area under the curve: AUC) was applied and demonstrated that these seizure forecasts performed better than chance (i.e. shuffled seizure times). Multiday seizure cycles are therefore similar in paediatric and adult cohorts, and this study provides the first validation of cycle-based seizure risk forecasting models as a promising approach for paediatric epilepsy.

## Introduction

Epileptic seizure times exhibit long-memory dynamics (1) and cyclic patterns (referred to as seizure cycles), following circadian, multiday, or even seasonal rhythms (2). Similar rhythms have also been observed in interictal epileptic activity (3,4), with seizures locked to particular phases of a cycle (4,5). At present, the underlying cause of these rhythms remains unknown, however, characterising seizure cycles could have valuable applications in managing and treating epilepsy. In adults, multiday seizure cycles can be used to develop forecasts of seizure risk days into the future (2). However, while multiday seizure cycles have been documented extensively in adults (6), substantive characterisation of these cycles and whether it can be applied within a forecasting framework is lacking in paediatric epilepsy. Therefore, this study aims to 1. identify and compare similar patterns of multiday seizure cycles in a paediatric cohort, and 2. assess whether these multiday seizure cycles can be applied to generate a personalised forecast of seizure likelihood within a paediatric cohort.

In order to measure seizure cycles, reliable long-term records of seizure times are required. Chronic EEG recording devices provide the most accurate method of seizure detection, yet these devices are invasive and rarely implanted in paediatric patients. However, evidence from intracranial EEG recordings suggests self-reported seizures can also be used to monitor multiday cycles (6,7), providing a non-invasive alternative whereby patients (or caregivers) can report seizures on mobile devices. Multiday cycles extracted from self-reported seizure times have been identified in adults across different seizure types and sexes (8), however, there is limited research characterising epileptic rhythms in children.

One of the first studies to identify seizure cycles in epilepsy demonstrated this phenomenon in a child, with an example of an approximately monthly cycle in an 11-year-old male (9). While this pioneering work demonstrated cases of multiday cycles at different ages (11, 20 and 40 years), there was no comparison of these seizure patterns in a larger paediatric population (9). Following this, short diurnal and sleep/wake seizure patterns were identified in infants (≤3 years), children (3 - 12 years) and adolescents (12 - 21 years), but multiday cycles were not explored (10). More recently, Wang et al (2022) conducted a large analysis of electronic seizure diaries to compare multiday cycle chronotypes across age groups (11). Various multiday cycles were estimated from individual seizure rates, which demonstrated that children aged 0 to 9 years were more likely to have tri-weekly and monthly cycles and less likely to have short, 2- to 4-day cycles (11). A significant benefit of the cyclic nature of the multiday patterns is the potential to project these cycles forward and estimate future seizure likelihood (7). While this approach is well-established within adult epilepsy, there are currently no studies which have employed a cycle-based approach to generate personalised seizure forecasts within a paediatric cohort. As such, the present study builds on previous findings by implementing a cycle-based approach drawn from seizure risk forecasting models (7,12). This is achieved by identifying and comparing multiday seizure cycles between paediatric and adult cohorts with epilepsy, which is then followed by the first validation of multiday seizure forecasting within a paediatric cohort.

## Materials & Methods

### Data

This study was approved by St Vincent’s Hospital Melbourne Human Research Ethics Committee (HREC 165.19). Participants with an epilepsy diagnosis confirmed on video-EEG monitoring were recruited to this study. Participants’ electronic seizure diaries were accessed via a cloud platform from a mobile epilepsy management app (Seer Medical). Participants with more than 20 seizures (excluding seizures recorded in the same hour) and diary length of at least 28 days were included for analysis. Those with more than 120 seizures reported per month were excluded. Participants’ age was determined as the age at the date of diary creation.

### Seizure cycles

Circadian (0.5 – 1.5 days) and multiday (2 – 32 days) seizure cycles were identified using the synchronisation index (SI, also called the phase-locking value) as previously described (2). In brief, the SI value measures the synchronisation of seizure times to an underlying cycle, where an SI value of 0 indicates no synchrony and 1 indicates perfect synchrony. The SI was computed for a range of cycle periods from 0.5 to 32 days (in increments of 0.5-days for circadian and 1-day for multiday). For each participant, the longest cycle period considered for analysis was 32 days or one quarter of the diary duration (whichever was less), where the diary duration was the number of days from diary creation to the last reported seizure (excluding seizure free periods greater than 50 days). This ensured that we captured at least 4 cycles of consistent diary use. To group cycles of interest, k-means clustering was performed on the cycle periods of all peak SI values, where peaks were selected at periods with a SI value greater than 0.3 (‘moderate synchronisation’ (2)). Centroids were initialised using the k-means++ method and the optimal number of clusters (from 1 to 10) were chosen using the sum of squared error and silhouette coefficient.

### Seizure forecasts

A probabilistic framework for estimating seizure likelihood was applied (described previously by Karoly et al., 2020). The strongest circadian “fast” (0.5 - 1.5 days) and multiday “slow” (2 - 32 days) cycles were extracted according to their SI values. For these fast and slow cycles, a Von Mises distribution of seizure risk was constructed based on past seizure occurrence. Retrospectively, seizure likelihoods for individual cycles were generated by mapping the Von Mises distribution onto the cycles in the time domain. Individual seizure likelihoods for each cycle were then combined using a log-odds model to produce a combined seizure likelihood value for every hour. Combined seizure likelihood values were used to calculate AUC-ROC scores for each individual.

### Statistics

Significant seizure cycles were identified using the Rayleigh test for periodicity (*p* < 0.05) with a Bonferroni correction for multiple comparisons. The distribution of the maximum SI value within each cluster was used to compare cycle strength between paediatric (<18 years) and adult (≥18 years) participants. The non-parametric Wilcoxon rank-sum test was used to test the null hypothesis that sum of the ranks did not differ between the two groups. Likewise, the Kolmogrov-Smirnov test was used to test whether the samples were drawn from populations with the same distribution. For both tests, the Benjamini Hochberg false discovery rate (FDR) correction was applied to address multiple corrections (with both raw p-values and adjusted values presented). A less conservative correction was specifically chosen (relative to Bonferroni) as we are examining ‘non-significance’ and it reduces the possibility of a type II error (false-negative). For all statistical tests, a *p*-value of <0.05 was considered statistically significant. Performance in seizure forecasting was assessed using the receiver operating characteristic (area under the curve: AUC) to measure forecasting performance compared to chance level AUC (i,e, shuffled seizure times). All analyses were conducted using Python (version 3.9.12).

## Results

### Patient cohort

From 1130 app users, 325 individuals (71 paediatric) met the inclusion criteria. Participants were split into paediatric <18 years (M = 10.7y, SD = 4.1y) and adult ≥18 years (M = 38.7y, SD = 15.4y) cohorts. From participants that specified their sex, there were 188 females (paediatric = 37) and 99 males (paediatric = 25). A total of 34,402 seizures were reported (paediatric: M = 101, SD = 103, adult: M = 107, SD = 156). The paediatric cohort reported an average of 22.1 (SD = 27.6) seizures per month, while adults reported 16.1 (SD = 19.4) seizures per month. Video-EEG monitoring provided evidence of 160 cases of focal (paediatric = 19) and 60 cases of generalised (paediatric = 32) seizures. The remainder of cases did not show clear evidence for either focal or generalised epilepsy (but did have evidence of epilepsy diagnosis as confirmed by video-EEG monitoring). Diary duration ranged from 14 - 1097 days (paediatric: M = 209, SD = 199, adult: M = 228, SD = 222). At the time of monitoring, 228 participants (paediatric = 44) were taking at least one anti-seizure medication.

### Characterisation of multiday cycles

Clusters (n = 1, 2,…,10) were compared and three multiday cycle groups were selected based on optimising the sum of square error and silhouette coefficient (Supplementary Figure 1). Seizure cycle groups were evaluated across circadian (0.5 - 1.5 days) and multiday seizure cycles, grouped according to the three clusters centred at 7.7 (2 – 12), 17.6 (13 – 22) and 27.3 (23 – 32) days (i.e. about-weekly, about-fortnightly and about-monthly cycles). Figure 1 shows the proportion of paediatric and adult cases with significant seizure cycles in each group. None of the proportions were significantly different after correcting for multiple comparisons across the four cycle groups (*p* > 0.05 using a standard z-test with Bonferroni correction).

**Figure 1.**
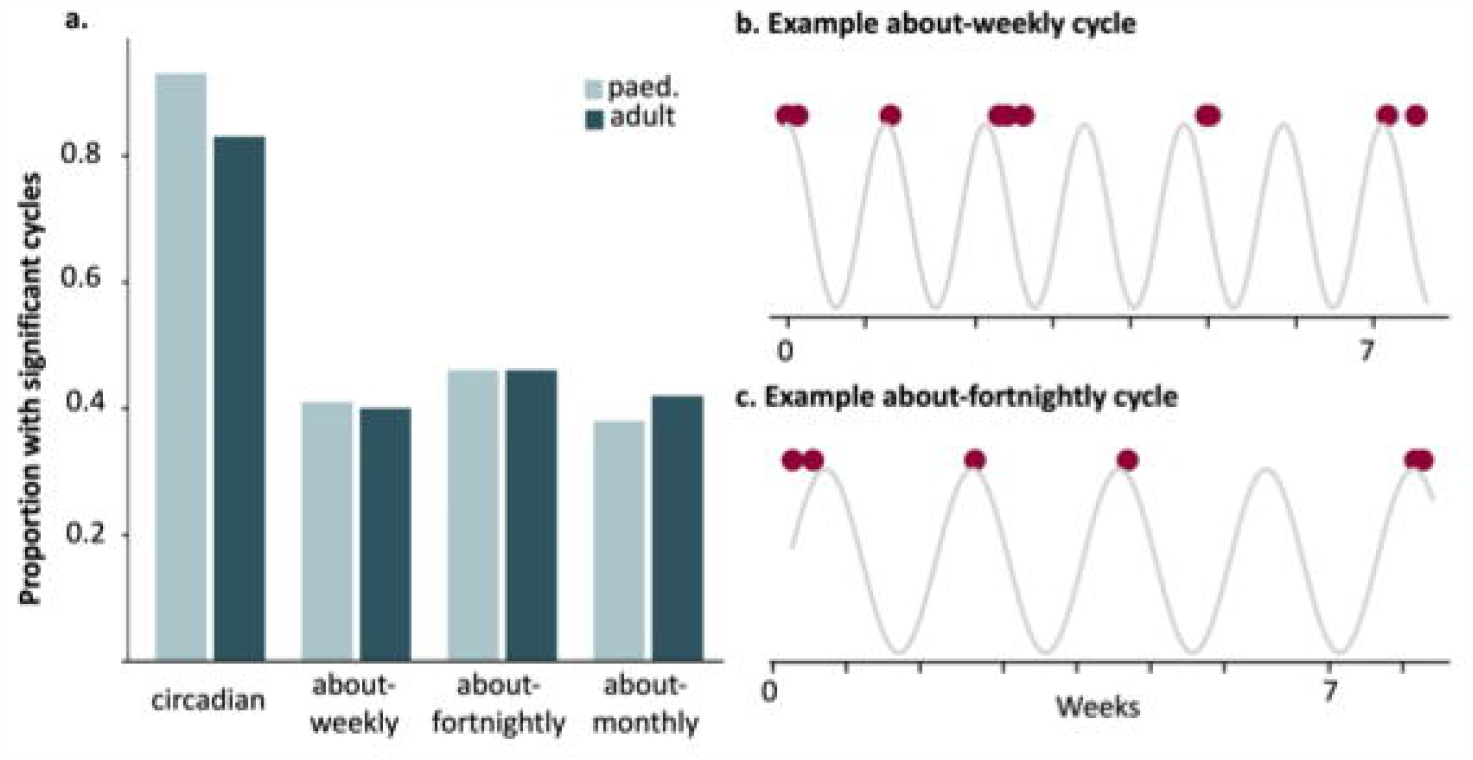
Significant seizure cycles. **a**. Proportion of paediatric and adult cohorts with significant seizure cycles (Rayleigh test, *p*< 0.05 with Bonferroni correction for multiple comparisons across the 4 cycle groups). Proportions were compared between paediatric and adult cohorts using a standard z-test (*p* = 0.04 for circadian, *p* = 0.87 for about-weekly, *p* = 0.99 for about-fortnightly, *p* = 0.63 for about-monthly) **b**. Example showing a 9yo with a 9-day (about-weekly) cycle (SI = 0.4) **c**. Example showing a 14yo with a 15-day (about-fortnightly) cycle (SI = 0.3). Red dots denote individual seizures in Figure 1b and c.

### Characterisation of cycle strength

Figure 2 shows the overall distributions in cycle strength (SI values) between paediatric and adult cohorts across circadian and multiday cycles. There were no significant differences between paediatric or adult distributions for any circadian or multiday cycle group.

**Figure 2.**
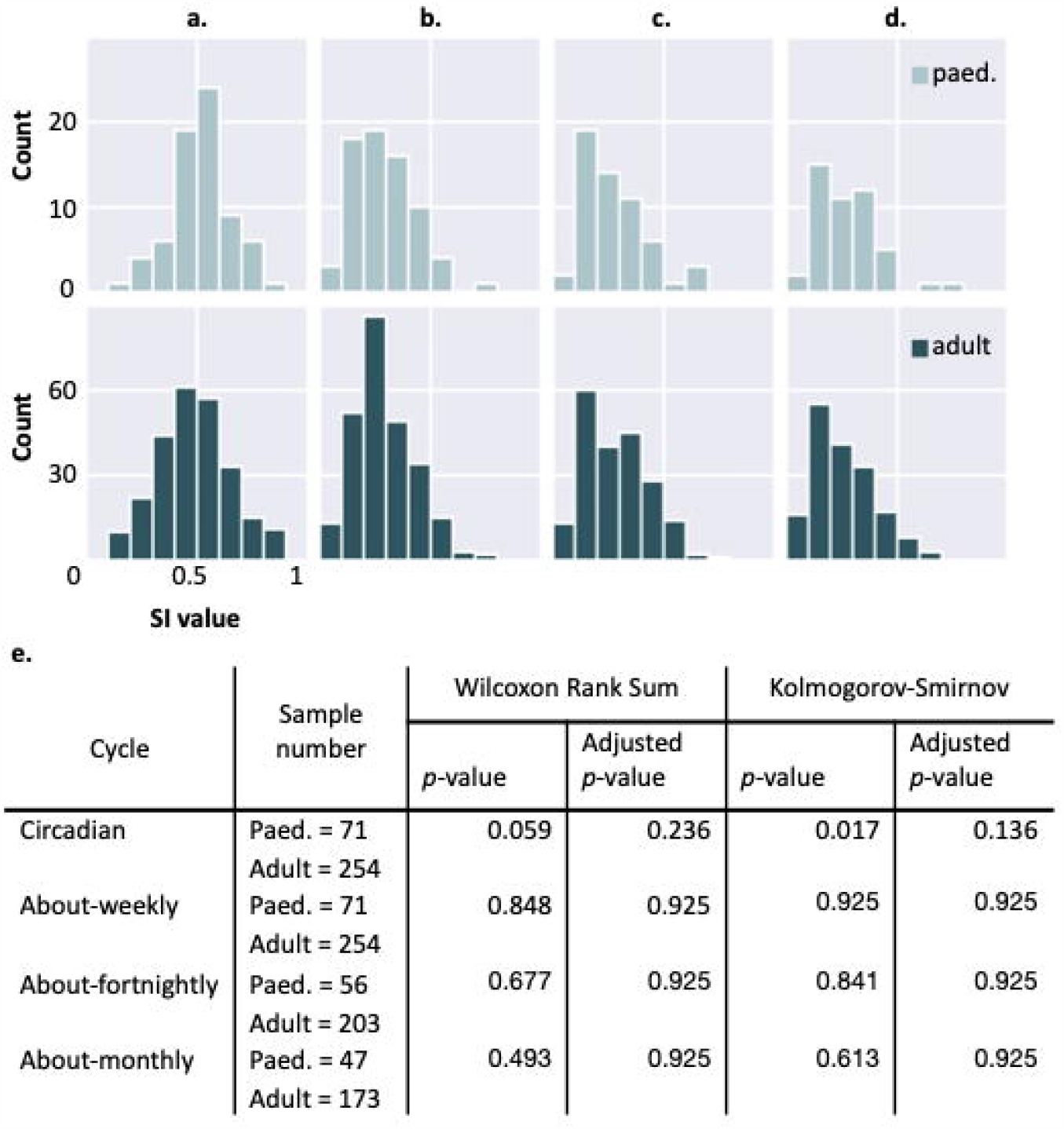
Distribution of the maximum SI values in each cycle group for paediatric and adult cohorts, **a**. circadian (0.5 - 1.5 days) **b**. about-weekly (2 - 12 days) **c**. about-fortnightly (13 - 22 days) **d**. about-monthly (23 - 32 days), and **e**. Statistical tests with adjusted p-values (computed using the Benjamini-Hochberg method), comparing the distribution of SI values between paediatric and adult cohorts for each cycle group.

Cycle strength distributions were also compared between paediatric and adult cohorts according to paediatric age (greater or less than 13-years-old), sex, epilepsy-type and medication-usage (Supplementary Figures 2, 3, 4 & 5 respectively), with no significant differences found for any circadian or multiday cycle groups (Supplementary Tables 1, 2, 3 & 4).

### Seizure forecasting performance

Across the paediatric cohort, the average AUC score (based on micro-average) was 0.77 with a range for the individual AUC scores between 0.64 to 0.91. To assess the accuracy of seizure forecasting performance, 95% confidence intervals were computed for the hourly AUC and compared to AUC for chance performance. AUC greater than 0.5 reflects performance above chance level. Figure 3. shows all individuals within the paediatric cohort presented with AUC values significantly better than chance, with non-overlapping confidence intervals.

**Figure 3.**
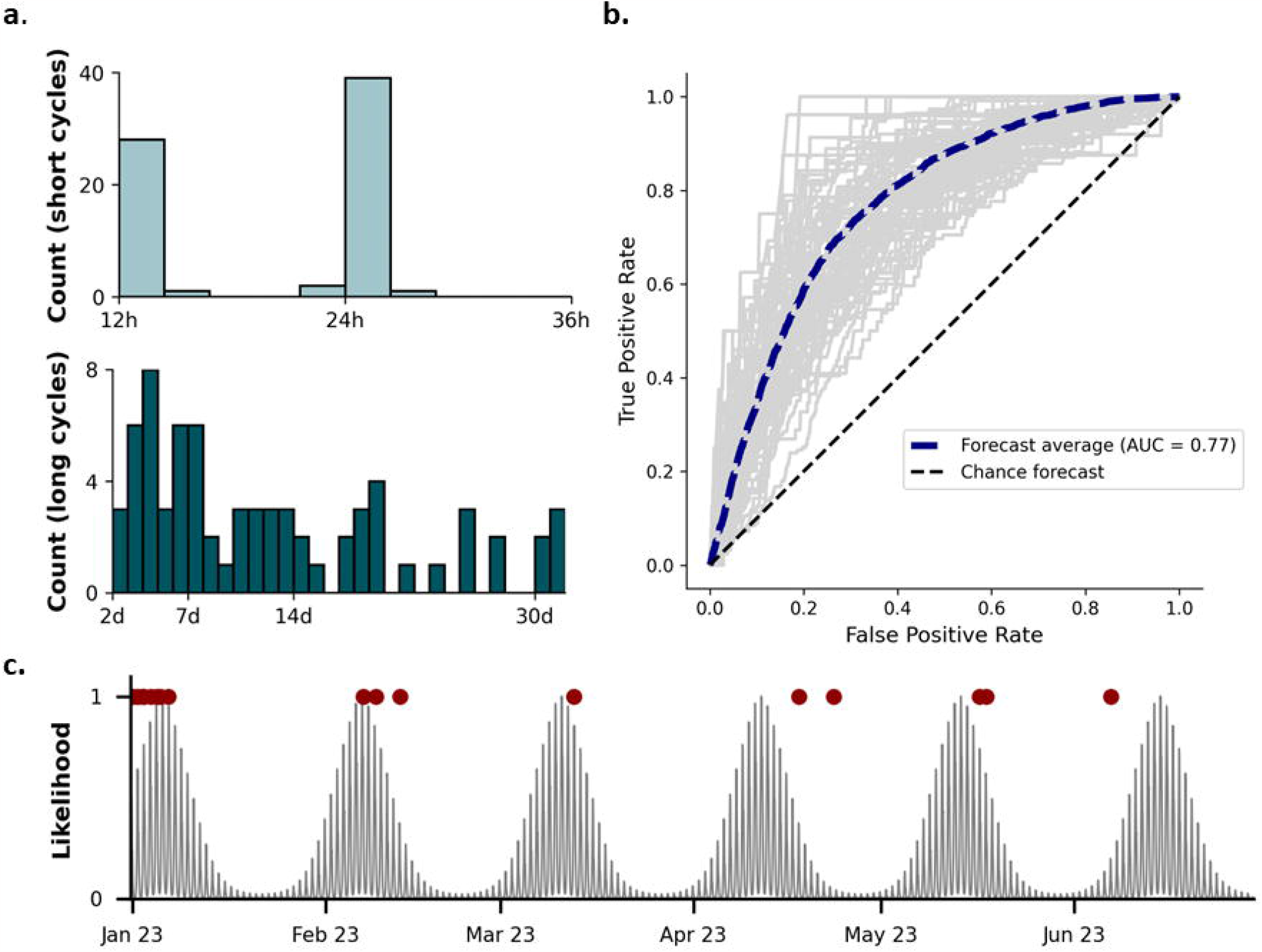
Receiver operating characteristic curves for seizure forecasting in paediatric cohort, **a**. histogram of the count of 1. short “circadian” cycles (above) with bar width of 2.4 hours and 2. long “multiday” cycles (below) with bar width of 1 day, these were the significant multiday cycles which formed the basis of the seizure forecast, **b**. receiver operating curve (ROC) with the AUC values for each of the participants presented as grey lines, the micro-average of all participant AUC values was calculated and presented as blue line (micro-average was applied given there were hours with seizures and hours without seizures included), the AUC of chance (i.e. based on shuffled seizure times) was presented in the diagonal black line. This demonstrates clearly that seizure forecasting within the paediatric cohort performed better than chance, **c**. provides an example forecast extracted from the paediatric cohort, the participant had a 24h circadian rhythm and a 32d about-monthly rhythm. Red dots mark the self-reported seizures and grey lines reflect the seizure likelihood which is extracted from the combined short “circadian” and long “multiday” cycles.

## Discussion

This study identified multiday seizure cycles in a paediatric cohort with confirmed epilepsy and provided the first evidence of the efficacy of cyclic-based seizure forecasting within a paediatric cohort. Understanding the presentation of these multiday cycles across different age groups may help to uncover the underlying mechanisms of action. The current study found no significant differences in the prevalence (Fig 1) or strength (Fig 2) of about-weekly, about-fortnightly, or about-monthly cycles between adult and paediatric cases, suggesting the existence of multiday cycles despite age-related factors. A recent study showed children (0 to 9 years) were more likely to have longer tri-weekly and monthly cycles and less likely to have short 2- to 4-day cycles than adults (11). The current study identified a similar trend, whereby paediatric participants showed slightly more monthly cycles, although the difference was not significant (Fig 1). The prevalence of monthly cycles in paediatric cohorts further reduces the likelihood that monthly seizure cycles are purely driven by hormonal changes associated with the menstrual cycle (13). In addition, the strength of monthly (and other) multiday cycle groups was similar between males and females at all age groups (Supplementary Fig 3, Supplementary Table 2). These findings, in conjunction with previous work (6,8,9), support the hypothesis that multiday seizure cycles are widespread, regardless of age or sex.

With regards to epilepsy type, the majority of studies into seizure forecasting (in adults) has been focused on individuals with focal epilepsy and there is very limited research into the efficacy of these techniques for generalised epilepsies. Our findings suggest comparable multiday cycles, in both prevalence and strength, between focal and generalised epilepsy, and this finding was consistent across both the adult and paediatric cohort. However, these findings are preliminary, therefore, further studies are required to explore the applicability of this approach across different types of epilepsy. In terms of medication effects, early studies suggested that while anti-seizure medication can dampen the number of seizures, the underlying seizure cycles appear to persist (9). Our findings support this hypothesis, as there were no significant differences in the prevalence or strength of cycles between those who were medicated with anti-seizure medications as compared to those who were not medicated, and this was consistent for both adults and paediatric cohorts.

Multiday cycles have important clinical applications in forecasting seizure risk, and the current study deployed a cycle-based approach that has been validated for prospective seizure risk forecasting in adults (12). While other methods have been explored to measure cycles, not all measures can be used to prospectively forecast future seizure risk. For example, Wang et al (2022) compared multiday seizure cycles in children using a Bayesian model based on current seizure rates; however, this model has not yet been explored to project future seizure risk. Other forecasting approaches have used machine learning, rather than cycles, to forecast seizure risk. Meisel et al (2020), showed that seizure forecasting using a wearable device was feasible for half of a paediatric cohort (n = 69), although recording duration was limited to several days (15). Goldenholz et al (2020) used a deep learning model with long-term seizure diaries to forecast 24-hour seizure risk in a large cohort (n = 5149) with median age of 17 (16), although performance was not explicitly assessed in children. This study is the first to assess the possibility of applying these cyclic-based seizure forecasting techniques in a paediatric cohort. Findings from the current study demonstrated that seizure forecasting models performed above chance and suggest that paediatric cohorts could benefit from cycle-based seizure risk forecasting, which can be used for tracking individual seizure risk (7,12) and scheduling epilepsy monitoring (17). Seizure risk forecasts could also improve the quality of life of children and caregivers, allowing them to gain more control over day-to-day activities, or by scheduling medication, monitoring/imaging studies, and other treatments. Further work is needed to validate the performance of a prospective risk forecasting model in a paediatric cohort, to ensure that this cohort would in fact benefit from such forecasts.

This study had several limitations. Self-reported seizure diaries have known issues with both under and over-reporting, and in the current study, reporting may have been further impacted by variations in whether patients or caregivers, or a combination of both, were documenting seizures. Despite these limitations, seizure cycles can still be detected (8) and may align to underlying cycles of chronic EEG in adults (6,7). Further cycle validation is challenging for paediatric cohorts because chronic implants in paediatric epilepsy are rare. The study was also limited by the small sample of paediatric diaries with sufficient duration to investigate longer rhythms (e.g. circannual) that have been found in a small proportion of adults with epilepsy (6). To capture longer cycles, seizure diaries spanning multiple years would be necessary, and may become possible in future studies with the ongoing use of electronic seizure diaries. Finally, multiday seizure cycles were extracted from a general paediatric cohort with epilepsy. However, as this was a preliminary study with a small sample size, we suggest larger studies should be conducted to further probe the efficacy of seizure forecasting techniques across various subgroups, such as epilepsy syndrome, age of onset, or seizure type.

## Conclusion

This study identified both circadian and multiday (about-weekly, about-fortnightly and about-monthly) seizure cycles in paediatric epilepsy that were comparable in prevalence and strength to those in adults. The cycle-based approach used in the current study has previously been validated for seizure risk forecasting in adults (7,12), and this forecasting approach was applied to the paediatric cohort, leading to the first validation of seizure forecasting for paediatric epilepsy. Therefore, the current findings demonstrate the feasibility of translating seizure risk forecasting technology for paediatric epilepsy populations, with potential benefits to their treatment outcomes and mental wellbeing.

## Data Availability

All data produced in the present study are available upon reasonable request to the authors.

## Acknowledgements/Funding

This research was supported by NHMRC Investigator Grant (1178220), My Seizure Gauge Grant (Epilepsy Foundation of America) and the BioMedTech Horizons 3 program (initiative of MTPConnect). Funding partners were not involved in the study design, collection, analysis, interpretation of data, the writing of this article or the decision to submit it for publication.

## CRediT (Contribution Roles Taxonomy)

H.K., P.J.K. and J.NF were involved in the conceptualization, formal analysis, methodology and writing (original draft preparation) of the study. R.E.S., E.S.N., D.F., M.J.C, W.D. and D.E. contributed to the conceptualization, methodology and writing (review and editing) of the manuscript.

## Ethical Publication Statement

The present study was approved by St Vincent’s Hospital Melbourne Human Research Ethics Committee (HREC 165.19).

