## Supplementary File for "Seizure cycles in paediatric epilepsy"

### Supplementary Figures

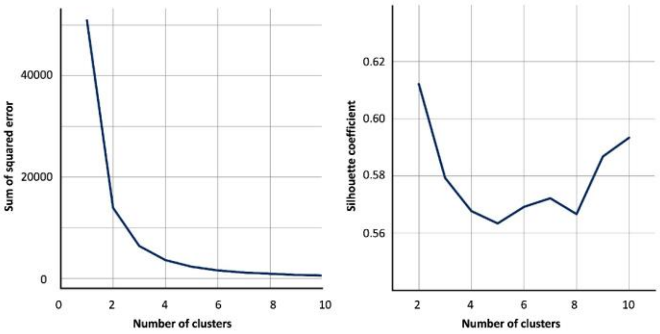


**Supplementary figure 1. Number of multi-day cycle groups determined by k-means clustering.** The optimal number of clusters (k) was identified by minimising the **a.** Sum of squared error at the elbow point (k = 3), and maximising the **b.** Silhouette coefficient.

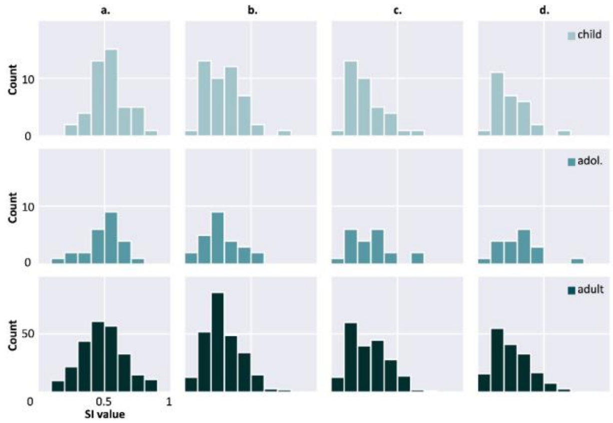


**Supplementary figure 2. Age subgroups.** Distribution of the maximum SI values in each cycle group for child (<13 years), adolescent (13 - 17 years) and adult (≥18 years) cohorts, **a.** circadian (0.5 - 1.5 days) **b.** about-weekly (2 - 12 days) **c.** about-fortnightly (13 - 22 days) and **d.** about-monthly (23 - 32 days)

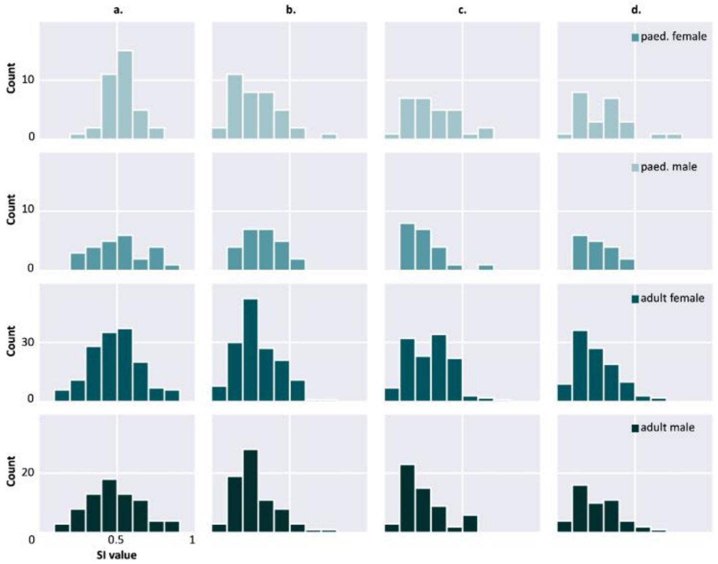


**Supplementary figure 3. Gender subgroups.** Distribution of the maximum SI values in each cycle group for paediatric female, paediatric male, adult female and adult male subgroups, **a.** circadian (0.5 - 1.5 days) **b.** about-weekly (2 - 12 days) **c.** about-fortnightly (13 - 22 days) and **d.** about-monthly (23 - 32 days)

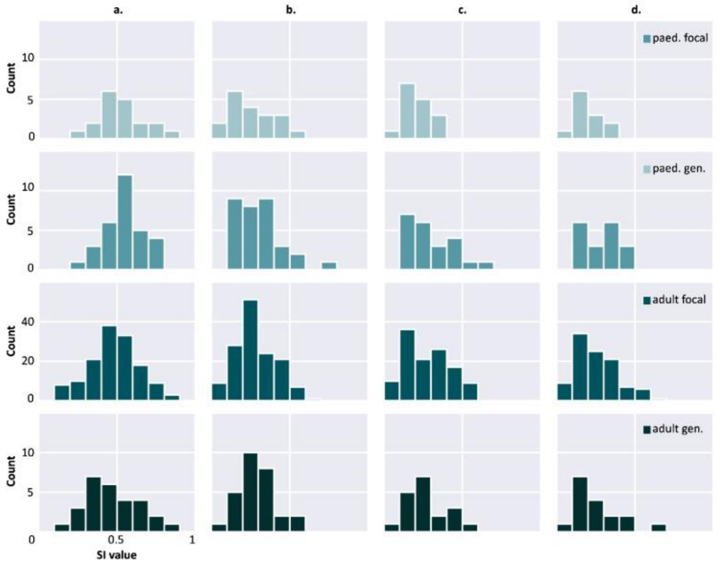


**Supplementary figure 4. Epilepsy type.** Distribution of the maximum SI values in each cycle group for paediatric focal, paediatric generalised, adult focal and adult generalised subgroups, **a.** circadian (0.5 - 1.5 days) **b.** about-weekly (2 - 12 days) **c.** about-fortnightly (13 - 22 days) and **d.** about-monthly (23 - 32 days)

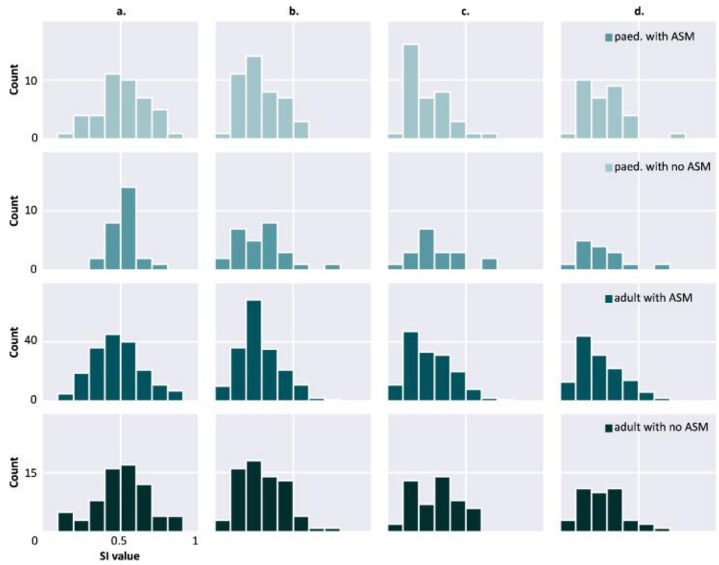


**Supplementary figure 5. Medication subgroups.** Distribution of the maximum SI values in each cycle group for paediatric with ASM, paediatric with no ASM, adult with ASM and adult with no ASM subgroups, **a.** circadian (0.5 - 1.5 days) **b.** about-weekly (2 - 12 days) **c.** about-fortnightly (13 - 22 days) and **d.** about-monthly (23 - 32 days)

### Supplementary Tables

| Cycle group | Comparison (n) | | Wilcoxon Rank Sum | | Kolmogorov-Smirnov | |
|---|---|---|---|---|---|---|
| | | | *p*-value | Adjusted *p*-value | *p*-value | Adjusted *p*-value |
| Circadian | Child (46) | Adolescent (25) | 0.531 | 0.974 | 0.957 | 0.974 |
| | Child (46) | Adult (254) | 0.046 | 0.552 | 0.023 | 0.552 |
| | Adolescent (25) | Adult (254) | 0.533 | 0.974 | 0.510 | 0.974 |
| About-weekly | Child (46) | Adolescent (25) | 0.876 | 0.974 | 0.948 | 0.974 |
| | Child (46) | Adult (254) | 0.954 | 0.974 | 0.907 | 0.974 |
| | Adolescent (25) | Adult (254) | 0.789 | 0.974 | 0.931 | 0.974 |
| About-fortnightly | Child (35) | Adolescent (21) | 0.548 | 0.974 | 0.667 | 0.974 |
| | Child (35) | Adult (203) | 0.490 | 0.974 | 0.477 | 0.974 |
| | Adolescent (21) | Adult (203) | 0.853 | 0.974 | 0.974 | 0.974 |
| About-monthly | Child (28) | Adolescent (19) | 0.165 | 0.712 | 0.106 | 0.712 |
| | Child (28) | Adult (173) | 0.878 | 0.974 | 0.736 | 0.974 |
| | Adolescent (19) | Adult (173) | 0.178 | 0.712 | 0.139 | 0.712 |

**Supplementary table 1. Age subgroups.** Statistical tests with adjusted *p*-values (computed using the Benjamini-Hochberg method) comparing the distribution of the maximum SI values in each cycle group for child (<13 years), adolescent (13 - 17 years) and adult (≥18 years) cohorts.

| Cycle group | Comparison (n) | | Wilcoxon Rank Sum | | Kolmogorov-Smirnov | |
|---|---|---|---|---|---|---|
| | | | *p*-value | Adjusted *p*-value | *p*-value | Adjusted *p*-value |
| Circadian | Paed female (37) | Paed male (25) | 0.514 | 0.847 | 0.508 | 0.847 |
| | Paed female (37) | Adult female (151) | 0.061 | 0.448 | 0.034 | 0.416 |
| | Paed male (25) | Adult male (74) | 0.425 | 0.847 | 0.794 | 0.847 |
| | Adult female (151) | Adult male (74) | 0.790 | 0.847 | 0.772 | 0.847 |
| About-weekly | Paed female (37) | Paed male (25) | 0.305 | 0.751 | 0.475 | 0.847 |
| | Paed female (37) | Adult female (151) | 0.720 | 0.847 | 0.666 | 0.847 |
| | Paed male (25) | Adult male (74) | 0.070 | 0.448 | 0.120 | 0.549 |
| | Adult female (151) | Adult male (74) | 0.179 | 0.643 | 0.116 | 0.589 |
| About-fortnightly | Paed female (28) | Paed male (21) | 0.210 | 0.643 | 0.241 | 0.643 |
| | Paed female (28) | Adult female (124) | 0.628 | 0.847 | 0.863 | 0.863 |
| | Paed male (21) | Adult male (58) | 0.610 | 0.847 | 0.770 | 0.847 |
| | Adult female (124) | Adult male (58) | 0.039 | 0.416 | 0.015 | 0.416 |
| About-monthly | Paed female (24) | Paed male (17) | 0.427 | 0.847 | 0.576 | 0.847 |
| | Paed female (24) | Adult female (106) | 0.214 | 0.643 | 0.234 | 0.643 |
| | Paed male (17) | Adult male (48) | 0.743 | 0.847 | 0.766 | 0.847 |
| | Adult female (106) | Adult male (48) | 0.861 | 0.863 | 0.622 | 0.847 |

**Supplementary table 2. Gender subgroups.** Statistical tests with adjusted *p*-values (computed using the Benjamini-Hochberg method) comparing the distribution of maximum SI values for paediatric female, paediatric male, adult female and adult male subgroups.

| Cycle group | Comparison (n) | | Wilcoxon Rank Sum | | Kolmogorov-Smirnov | |
|---|---|---|---|---|---|---|
| | | | *p*-value | Adjusted *p*-value | *p*-value | Adjusted *p*-value |
| Circadian | Paed focal (19) | Paed generalised (32) | 0.360 | 0.683 | 0.496 | 0.794 |
| | Paed focal (19) | Adult focal (141) | 0.307 | 0.683 | 0.260 | 0.647 |
| | Paed generalised (32) | Adult generalised (28) | 0.025 | 0.371 | 0.033 | 0.371 |
| | Adult focal (141) | Adult generalised (28) | 0.675 | 0.864 | 0.822 | 0.974 |
| About-weekly | Paed focal (19) | Paed generalised (32) | 0.185 | 0.556 | 0.058 | 0.371 |
| | Paed focal (19) | Adult focal (141) | 0.325 | 0.682 | 0.384 | 0.682 |
| | Paed generalised (32) | Adult generalised (28) | 0.767 | 0.944 | 0.628 | 0.864 |
| | Adult focal (141) | Adult generalised (28) | 0.648 | 0.864 | 0.459 | 0.773 |
| About-fortnightly | Paed focal (16) | Paed generalised (22) | 0.098 | 0.523 | 0.132 | 0.528 |
| | Paed focal (16) | Adult focal (119) | 0.124 | 0.528 | 0.053 | 0.371 |
| | Paed generalised (22) | Adult generalised (19) | 0.583 | 0.864 | 0.857 | 0.979 |
| | Adult focal (119) | Adult generalised (19) | 0.988 | 0.988 | 0.942 | 0.980 |
| About-monthly | Paed focal (12) | Paed generalised (18) | 0.038 | 0.371 | 0.191 | 0.556 |
| | Paed focal (12) | Adult focal (103) | 0.188 | 0.556 | 0.263 | 0.647 |
| | Paed generalised (18) | Adult generalised (17) | 0.373 | 0.683 | 0.603 | 0.864 |
| | Adult focal (103) | Adult generalised (17) | 0.949 | 0.980 | 0.937 | 0.980 |

**Supplementary table 3. Seizure subgroups.** Statistical tests with adjusted *p*-values (computed using the Benjamini-Hochberg method) comparing the distribution of maximum SI values for paediatric focal, paediatric generalised, adult focal and adult generalised subgroups.

| Cycle group | Comparison (n) | | Wilcoxon Rank Sum | | Kolmogorov-Smirnov | |
|---|---|---|---|---|---|---|
| | | | *p*-value | Adjusted *p*-value | *p*-value | Adjusted *p*-value |
| Circadian | Paed with ASM (44) | Paed with no ASM (27) | 0.887 | 0.957 | 0.126 | 0.632 |
| | Paed with ASM (44) | Adult with ASM (184) | 0.075 | 0.632 | 0.183 | 0.651 |
| | Paed with no ASM (27) | Adult with no ASM (70) | 0.778 | 0.957 | 0.139 | 0.632 |
| | Adult with ASM (184) | Adult with no ASM (70) | 0.124 | 0.632 | 0.269 | 0.783 |
| About-weekly | Paed with ASM (44) | Paed with no ASM (27) | 0.776 | 0.957 | 0.922 | 0.957 |
| | Paed with ASM (44) | Adult with ASM (184) | 0.867 | 0.957 | 0.957 | 0.957 |
| | Paed with no ASM (27) | Adult with no ASM (70) | 0.760 | 0.957 | 0.851 | 0.957 |
| | Adult with ASM (184) | Adult with no ASM (70) | 0.361 | 0.825 | 0.298 | 0.795 |
| About-fortnightly | Paed with ASM (37) | Paed with no ASM (19) | 0.117 | 0.632 | 0.158 | 0.632 |
| | Paed with ASM (37) | Adult with ASM (153) | 0.479 | 0.957 | 0.616 | 0.957 |
| | Paed with no ASM (19) | Adult with no ASM (50) | 0.936 | 0.957 | 0.815 | 0.957 |
| | Adult with ASM (153) | Adult with no ASM (50) | 0.098 | 0.632 | 0.101 | 0.632 |
| About-monthly | Paed with ASM (32) | Paed with no ASM (15) | 0.819 | 0.957 | 0.872 | 0.957 |
| | Paed with ASM (32) | Adult with ASM (132) | 0.348 | 0.825 | 0.572 | 0.957 |
| | Paed with no ASM (15) | Adult with no ASM (41) | 0.677 | 0.957 | 0.610 | 0.957 |
| | Adult with ASM (132) | Adult with no ASM (41) | 0.264 | 0.783 | 0.482 | 0.957 |

**Supplementary table 4. Medication subgroups.** Statistical tests with adjusted *p*-values (computed using the Benjamini-Hochberg method) comparing the distribution of maximum SI values for paediatric with ASM, paediatric with no ASM, adult with asm and adult with no ASM subgroups.
